# Burden of aortic aneurysm and its attributable risk factors in 204 countries and territories, 1990-2021 results from the Global Burden of Disease Study 2021

**DOI:** 10.1101/2025.06.20.25330025

**Authors:** Tong Jing, Quan Liu, Dongliang Chen, Xunli Zhang, Yueyun Zhou, Jian Li, Shengchen Liu, Wen Chen, Fuhua Huang

## Abstract

**Objective:** To assess the global, regional, and national burden of aortic aneurysm (AA) and its risk factor attributions by age, sex, and sociodemographic index (SDI) from 1990 to 2021.

**Design:** Cross-sectional analysis of epidemiological trends using the Global Burden of Diseases, Injuries, and Risk Factors Study (GBD) 2021 data.

**Main outcome measures:** Age-standardised mortality rate (ASMR), disability-adjusted life years (DALYs), and attributable risk factors for AA.

**Methods:** Age-standardised rates and absolute counts of deaths and DALYs were calculated with 95% uncertainty intervals (UIs). Trends were analysed using annual percentage changes. Risk factor contributions were estimated via comparative risk assessment.

**Data sources:** Nationally representative mortality and morbidity data from 204 countries and territories, collated in the GBD 2021 study (1990–2021).

**Results:** In 2021, AA caused 153 900 deaths (ASMR: 1.9 per million, 26.7% decline since 1990) and 3.1 million DALYs (age-standardised DALY rate: 36.5 per 100 000, 25.1% decline since 1990).Armenia had the highest ASMR (9.2 per million) and DALY rate (192.8 per 100 000), while Saudi Arabia had the lowest (0.2 per million and 5.1 per 100 000). DALY rates peaked at 85-89 years in men (declining thereafter) but increased continuously with age in women (highest in ≥95 years). Age-standardised DALY rates exhibited a reversed V-shaped relationship with SDI, with higher burdens in low-to-middle SDI regions. Smoking (37.3%), hypertension (17.1%), and high BMI (8%) accounted for 62.4% of AA-related DALYs.

**Conclusions:** AA remains a critical public health challenge, disproportionately affecting low-SDI regions. Targeted interventions addressing smoking cessation, blood pressure control, and BMI regulation are essential to mitigate the burden. Policymakers should prioritise these strategies, particularly in socioeconomically disadvantaged populations.

## Introduction

Aortic aneurysm (AA) is a severe cardiovascular event characterized by abnormal dilation of the aorta, which can lead to life-threatening rupture if not timely diagnosed or treated. It frequently occurs in the thoracic and abdominal sections of the aorta, with the latter being much more prevalent than the former^1^. AA is typically asymptomatic until it reaches a critical size, posing challenges for early detection and significantly increasing the risk of fatal rupture^2^. The mortality rate associated with ruptured AA remains alarmingly high. One study indicates that approximately 80% of patients with ruptured abdominal AA succumb before receiving medical care if left untreated ^3^. AA ranks as the fifteenth leading cause of death among individuals aged 55 years old and older and the nineteenth leading cause of death across all age groups^4^. In recent years, shifts have been observed in the age-standardized rate of death (ASRD) from AA globally, decreasing from 2.70 per hundred thousand in 1990 to 2.21 per hundred thousand in 2019, with a reduction of approximately 17.9%. Notably, this decline was most pronounced in regions with a high Socio-Demographic Index (SDI)^5^. This trend may be influenced by advancements in imaging technology, an aging population, and changes in lifestyle ^6–8^. In the United States, improvements in diagnostic imaging have enhanced early detection rates, with reports indicating that AA rupture-related mortality decreased by approximately 50% between 2000 and 2020^9^. However, these advancements have primarily occurred in high-SDI regions. In contrast, there are the largest increases in ASRD in the low- and middle-SDI regions, particularly among men ^5^. Limited diagnostic resources in these countries underscore a significant disparity in outcomes.

The risk factors for AA include gender, smoking, hypertension, genetic susceptibility, and lifestyle factors, with age being one of the most critical determinants^7, 10, 11^. Men are at a notably higher risk than women, with studies indicating a prevalence of approximately 4 to 6% in men over 65 years old, compared to 1 to 2% in women^12^. Smoking is the leading modifiable risk factor, nearly doubling the risk of aneurysm compared to that of non-smokers^13^. Hypertension also plays a key role in increasing stress on the aortic wall, thereby promoting aneurysm formation and expansion^11^. Additionally, genetic predisposition, particularly among individuals with a family history of aneurysms-further elevates risk, underscoring the need for targeted screening in these populations ^10^.

Despite these advancements, limitations in current research hinder a comprehensive understanding of the global burden of AA. Many studies are regionally restricted, relying on data from certain countries, which may not capture the distinct epidemiological patterns present in low- and middle-income nations^14, 15^. Additionally, previously published GBD data lack information on theprevalence and incidence of AA, resulting in an absence of studies offering detailed time trends for incidence and prevalence of AA over extended periods^5, 16^. This restricts our ability to observe shifts in the disease burden across populations with diverse demographic and socioeconomic backgrounds. Addressing this data gap will require more granular examinations of AA prevalence, outcomes, and attributable risk factors across diverse populations to inform targeted interventions and improve global health outcomes.

The Global Burden of Disease (GBD) database, managed by the Institute for Health Metrics and Evaluation (IHME), offers a comprehensive and systematically collected dataset on the incidence, prevalence, mortality, and other key metrics for a wide range of diseases across 204 countries and territories. Utilizing standardized methods and advanced statistical models, this database facilitates robust comparisons across regions and populations. Our study reported the mortality, and disability-adjusted life years (DALYs) associated with AA from the GBD 2021 database across 204 countries and territories from 1990 to 2021, as well as attributable risk factors by age, sex, and Socio-Demographic Index (SDI). By integrating GBD data, our study aimed to fill the existing knowledge gaps, support evidence-based policy-making, and guide efficient resource allocation, as well as to inform strategies for reducing the burden of AA, particularly in resource-limited settings, contributing to global efforts to improve cardiovascular health.

## Materials and Methods

### Overview of the GBD 2021 Database

This study utilized data from the Global Burden of Disease (GBD) 2021 database, a comprehensive epidemiological resource compiled by the Institute for Health Metrics and Evaluation (IHME). The 2021 GBD study provided a detailed assessment of the health impacts of 369 diseases, injuries, and 88 risk factors across 204 countries and territories^17^. This analysis was based on the latest epidemiological data and enhanced standardization techniques. Data were downloaded from the Global Health Data Exchange query tool (http://ghdx.healthdata.org/gbd-results-tool) and categorized by region, sex, country, and risk factors. This study focused on outlining the burden of AA.

### Definition of Aortic Aneurysm

AA was defined in this study based that a condition where the abdominal or thoracic aorta is abnormally enlarged and weakened due to atherosclerosis, high blood pressure, changes in cellular or extracellular structure, or inflammation, which can lead to tearing or rupture of the blood vessels.

### Data Processing and Modeling Techniques

GBD 2021 applies stringent data processing and modeling techniques to compile and validate burden of disease estimates, utilizing advanced methods such as DisMod MR 2.1 and the Cause of Death Ensemble Model (CODEm)^18, 19^. CODEm integrates multiple data sources and assesses predictive validity by testing various model configurations. DisMod MR 2.1, a Bayesian meta-regression tool for disease modeling, enhances accuracy in estimating disease burden. In instances of data gaps, GBD addresses potential limitations through supplementary predictive modeling and expert consultations.

### Social and Demographic Index (SDI) for Comparative Analysis

To assess the impact of socioeconomic development on the burden of AA, this study utilized the Socio-Demographic Index (SDI). SDI is a composite metric ranging from 0 to 1 that combines income per capita, average educational attainment, and fertility rate among females under 25, reflecting a country’s level of social development^17^. In the GBD 2021 study, countries are categorized into five quintiles based on SDI (high, high-middle, middle, low-middle, and low) and 21 geographic regions, enabling an analysis of disease burden variations associated with different socioeconomic conditions^19^.

### Risk Factor Attribution and Comparative Assessment

The study examined several risk factors for AA, including smoking, diet low in fruits, diet high in sodium, diet low in vegetables, high body-mass index, and high systolic blood pressure. The GBD comparative risk assessment framework was used to estimate the fraction of DALYs and deaths attributable to each risk factor, applying population-attributable fractions to quantify their individual contributions to the total burden of AA.

## Results

### Global level

In 2021, 153.9 thousand deaths from AA were reported, with an age-standardized rate of 1.9 per million, a decrease of 26.7% since 1990. In 2021, the number of DALYs for AA globally was 3.1 million, with an age-standardized rate of 36.5 DALYs per hundred thousand, making a 25.1% of decrease since 1990 (table 1)

**Table 1.**
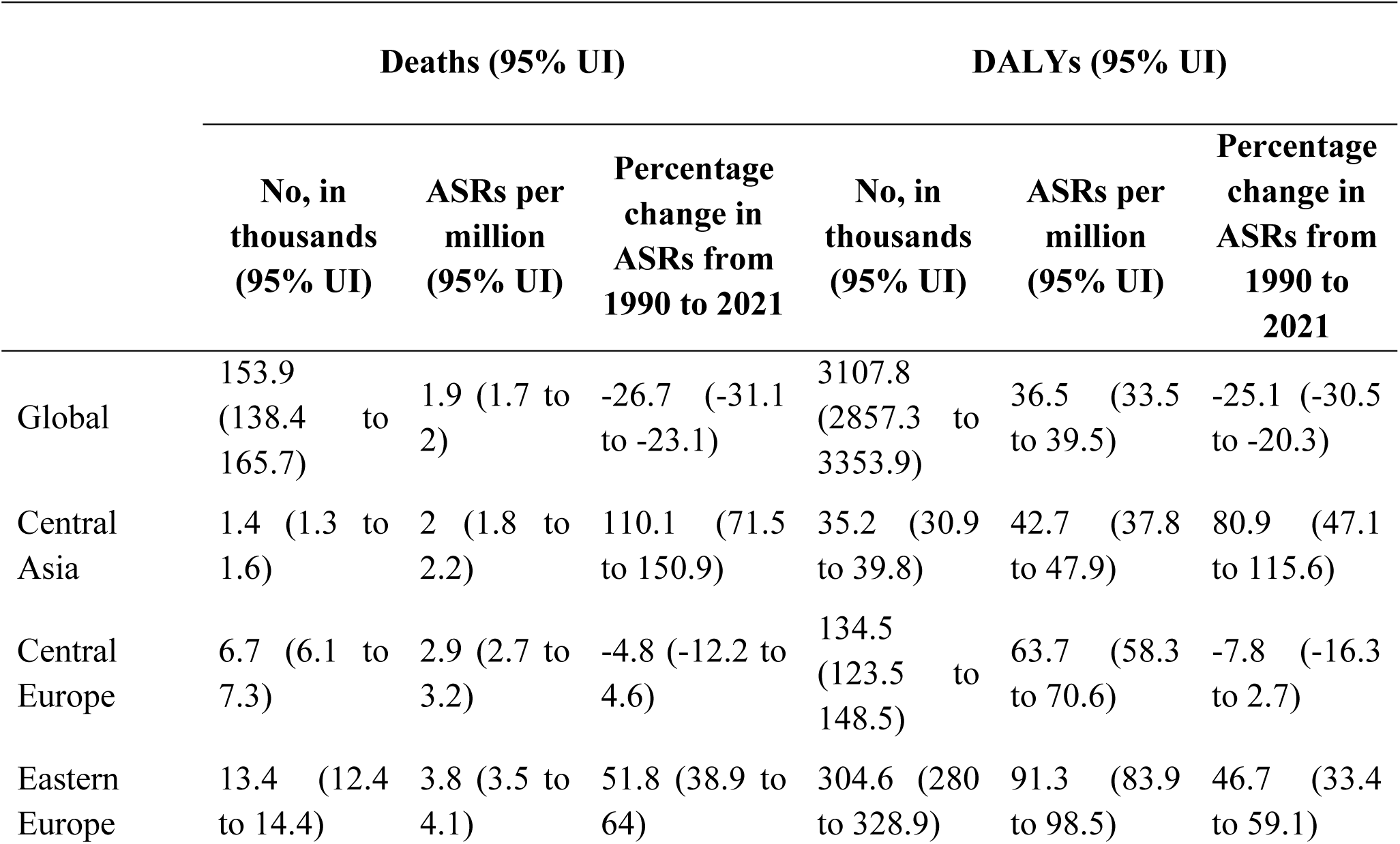
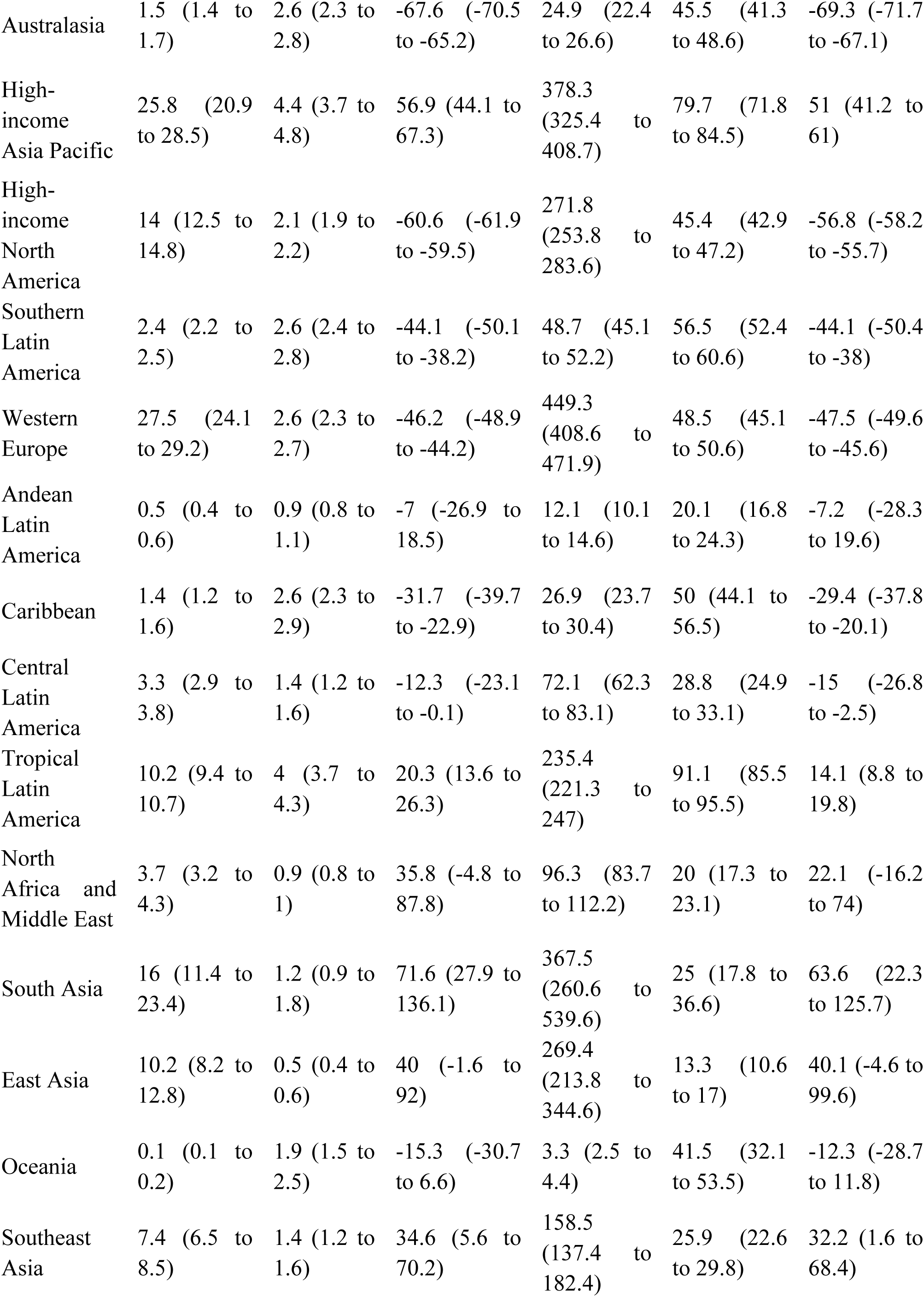
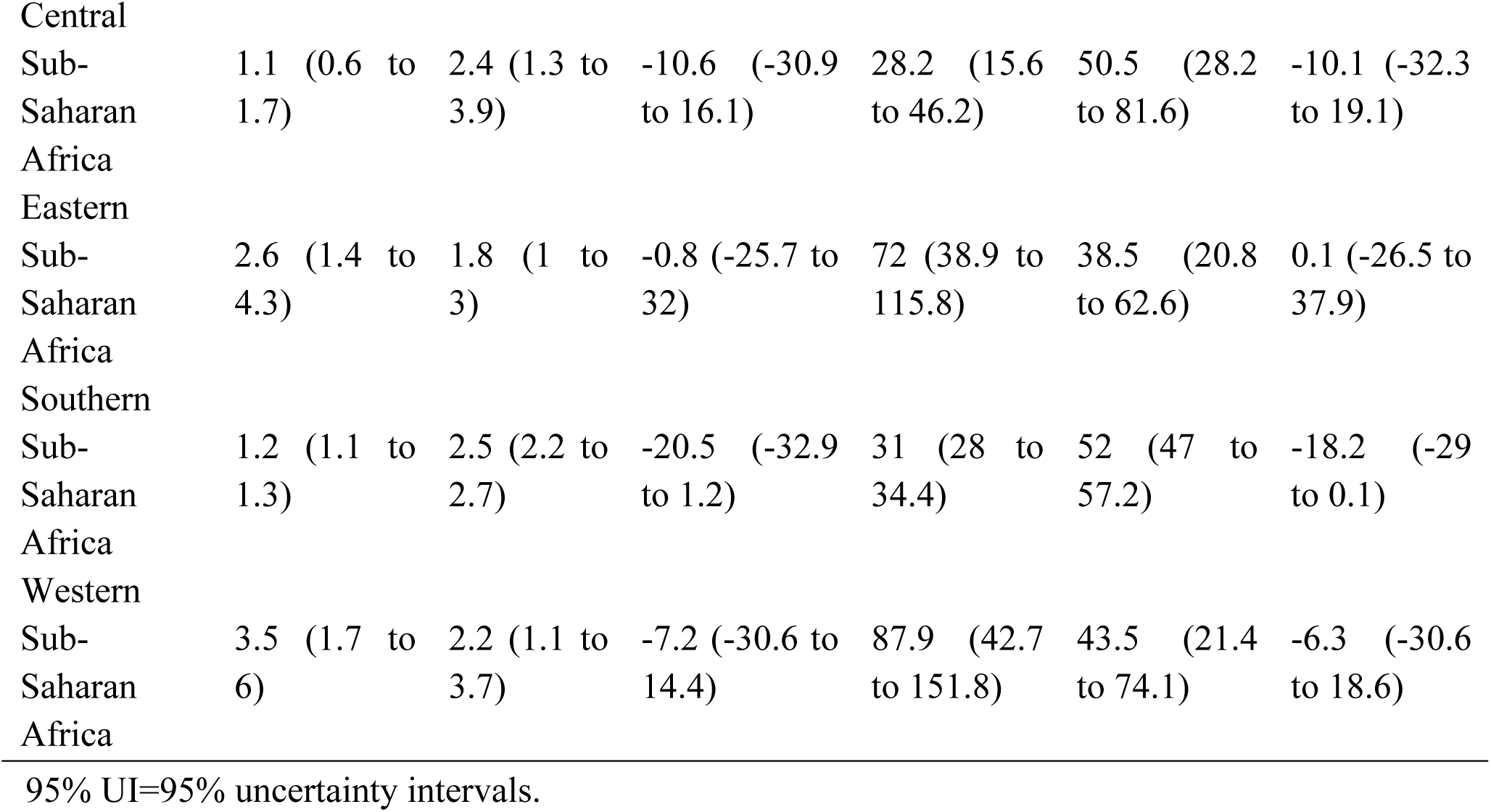
Deaths, and disability adjusted life years (DALYs) for AA in 2021, and percentage change in age standardized rates (ASRs) per hundred thousand, by Global Burden of Disease region, from 1990 to 2021 (generated from data available at https://ghdx.healthdata.org/gbd-results-tool)

### Regional level

In 2021, the highest age-standardized death rates for AA were observed in high-income Asia Pacific (4.4), Tropical Latin America (4), and Eastern Europe (3.8), whereas the lowest rates were found in North Africa and the Middle East (0.9), Andean Latin America (0.9), and East Asia (0.5) standardized (table 1). Eastern Europe (91.3), Tropical Latin America (91.1), and high-income Asia Pacific (79.7) had the highest age standardized DALY rates (per hundred thousand), whereas Andean Latin America (20.1), North Africa and Middle East (20), and East Asia (13.3) had the lowest (table 1). Figures S1 to S2 display the age standardized death, and DALY rates of AA, respectively, by sex in 2021 for all regions in the Global Burden of Disease study.

The largest increases in the age-standardized death rates for AA, from 1990 to 2021, were observed in Central Asia (110.1%), South Asia (71.6%), and high-income Asia Pacific (56.9%), while the most significant decreases were seen in Australasia (-67.6%), high-income North America (-60.6%), and Western Europe (-46.2%) (table 1). In the same period, the highest increases in age-standardized DALY rates were occurred in Central Asia (35.2%), South Asia (63.6%), high-income Asia Pacific (51%), with the largest decreases in Australasia (-69.3%), high-income North America (-56.8%), and Western Europe (-47.5%). Figures S3-S4 show the percentage changes in age-standardized death and DALY rates for AA by sex, respectively, from 1990 to 2021standardized.

The number of deaths caused by AA increased from 88.4 thousand in 1990 to 153.9 thousand in 2021, with the highest numbers of deaths in 2021 in Western Europe, high-income Asia Pacific, and South Asia (table S1). The number of DALYs due to AA increased from 1.9 million in 1990 to 3.1 million in 2021, with the highest numbers of DALYs in 2021 in Western Europe, high-income Asia Pacific, and South Asia (table S2).

### National level

Similarly, national age-standardized Death rates for AA in 2021 ranged from 0.2 to 9.2 per hundred thousand, standardized. The highest rates were observed in Armenia (9.2), Montenegro (8.7), and Nauru (6), whereas the lowest rates in Saudi Arabia (0.2), Tajikistan (0.3), and Nicaragua (0.3) (fig 1 and table S1). In 2021, the national age standardized DALY rate of AA ranged from 5.1 to 192.8 patients per hundred thousand. The highest rates were seen in Armenia (192.8), Montenegro (178.9), and Nauru (114.5) whereas the lowest rates in Saudi Arabia (5.1), Tajikistan (5.7), and Sri Lanka (6.7) (fig 2 and table S2).

The percentage change in the age-standardized death rate, from 1990 to 2021, differed noticeably between countries, with the largest increases in Uzbekistan (385.9%), Georgia (374.2%), and Morocco (231.7%). In contrast, Guam (-74.4%), Australia (-69%), and Canada (-65.7%) had the largest decreases (table S1). Over the same period, Georgia (357.4%), Uzbekistan (320.6%), and Sudan (209.4%) had the largest increases in the age standardized DALY rate, whereas the largest decreases were found in Australia (-70.6%), Bermuda (-66.6%), and Canada (-65.9%) (table S2).

### Age and sex patterns

In 2021, the global death rate of AA began to increase in the 30 to 34 age group, peaked in the oldest age group (≥95 years), and was higher in men across all age groups. Similarly, the highest number of deaths occurred in the 70 to 74 age group for men, then decreased with increasing age. Whereas for women, the number of deaths was the highest in the 80 to 84 age group. The number of deaths of AA was higher in men up the number of deaths was highest in the 80-84 to age of 85 to 89 years, but the number of deaths of AA was higher in women older than 89 years (fig. 3).

The global DALY rate for AA increased with age, peaking in the oldest age group (≥ 95 years old) for both sexes. This rate was consistently higher in men across all age groups. The number of DALYs peaked in the 65 to 69 age group for men, while it peaked in the 70 to 74 age group for women. DALYs were higher in men up to the 90 to 94 age group (fig. S5).

### Association with the sociodemographic index

At regional level, we found a reversed V-shaped association between the sociodemographic index and the age standardized DALY rate of AA, from 1990 to 2021. The age standardized DALY rate increased exponentially with increases in sociodemographic index, up to a sociodemographic index of about 0.7, before decreasing again. Central Sub-Saharan Africa, Western Sub-Saharan Africa, Oceania, Tropical Latin America, Southern Sub-Saharan Africa, Caribbean and Eastern Europe had higher than expected DALY rates, based on their sociodemographic index, from 1990 to 2021. In contrast, South Asia, North Africa and Middle East, Southeast Asia, East Asia, Central Latin America, Andean Latin America and Central Asia had lower than expected burdens from 1990 to 2021 (fig 4).

At the country level, in 2021, the burden of AA increased with increasing socioeconomic development up to a sociodemographic index of about 0.37, decreased up to a sociodemographic index of about 0.5 (fig S6), but then increased up to a sociodemographic index.

Countries and territories such as Armenia, Montenegro, Nauru and Russian Federation had much higher than expected burdens, whereas Afghanistan, Tajikistan, Sri Lanka and Saudi Arabia had much lower burdens than expected (fig S6).

### Risk factors

The proportion of DALYs due to AA that were attributable to individual risk factors differed across the Global Burden of Disease regions. Globally, smoking (37.3%), high systolic blood pressure (17.1%), and high body-mass index (8%) had the highest contributions to DALYs due to AA (fig. 5). The proportion of DALYs due to AA that were attributable to the smoking and high body-mass index was higher in men, whereas high systolic blood pressure was higher in women (fig S7 and S8), The proportion of DALYs due to AA attributable to individual risk factors also differed by age group. The proportion of attributable DALYs for smoking increased with age up to 40 to 44 years old and then decreased. Although the highest attributable DALYs for High systolic blood pressure was in the 65 to 69 age group, values did not differ significantly from other age groups. Moreover, the DALYs due to AA attributable to High body-mass index increased with age, and the highest proportion was found in the 60 to 64 age group (9.5%) (fig S9, S10 and S11) show the proportion of DALYs due to AA attributable to the individual risk factors for men and women, respectively.

## Discussion

In our study. we have provided up-to-date information on the death, DALY counts of AA from 1990 to 2021, and the age standardized rates across 204 countries and territories. Globally, AA accounted for 153.9 thousand deaths, and 3.1million DALYs in 2021. Although age-standardized mortality and DALY rates attributable to abdominal AA have decreased over the past three decades, the absolute number of cases has risen, likely as a result of population growth, aging, and increased life expectancy.

A study published 21 years ago indicated that from 2011 to 2015, the mortality rate due to abdominal AA decreased by 3.42% in males, while decreased by 2.61% in females.^20^ In our study, we found that from 1990 to 2021, the global age-standardized rate of DALY and mortality rate for AA declined by 25.1% and 26.7%, respectively. The observed discrepancies may be attributed to differences in the study periods and methodologies employed. Furthermore, due to methodological variations between the two datasets, direct comparison with the 2015 Global Burden of Disease study is challenging.^21^ A recent study has reported the burden of aneurysms from 1990 to 2019, and the overall findings regarding changes in mortality and DALY counts are consistent with those of our research.^5^

We attributed the decline in the burden of AA over time to several factors, including tobacco control measures, hypertension prevention and treatment initiatives, non-communicable disease programs, the establishment of chest pain centers in China, and aneurysm screening in European countries.^22, 23^ However, the Global Burden of Disease project relies on estimates and modeling processes rather than reporting actual data on the AA burden. This methodology may result in an underestimation of the true burden of AA, potentially leading to misguided policy decisions.

At the regional level, we observed an upward trend in both age-standardized mortality and DALY rates over the past 31 years in regions such as Central Asia, South Asia, High-income Asia Pacific, Eastern Europe, East Asia, North Africa and the Middle East, Southeast Asia, and Tropical Latin America. This trend aligns with previous studies^24, 25^. Several factors are likely contributing to this phenomenon, particularly the ongoing demographic aging in Asia, the prevalence of tobacco use, and hypertension. For instance, the high prevalence of hypertension in East Asia and the Pacific, South Asia, and Sub-Saharan Africa, coupled with limited investment in public health policies in these regions, may exacerbate the burden of cardiovascular diseases and other chronic conditions^26, 27^. Furthermore, shifts in smoking and dietary habits, particularly the widespread use of tobacco and the consumption of high-salt, high-fat diets, may further intensify the disease burden in these areas^28, 29^.

Conversely, in regions such as Eastern Sub-Saharan Africa, Central Europe, Andean Latin America, Western Sub-Saharan Africa, Central Sub-Saharan Africa, Central Latin America, Oceania, Southern Sub-Saharan Africa, the Caribbean, Southern Latin America, Western Europe, High-income North America, and Australasia, mortality and DALY rates have shown a downward trend. These changes likely reflect the effectiveness of diverse public health strategies, particularly improvements in disease prevention and treatment. For example, the declining trends observed in Western Europe and High-income North America may be closely linked to the recent advances in health promotion policies, the widespread adoption of disease screening, and improvements in public health infrastructure^30–32^.

From a policy perspective, we recommend the formulation of region-specific public health strategies. For areas where mortality and DALY rates continue to rise, particularly in Asia and Eastern Europe, greater emphasis should be placed on chronic disease intervention, with a focus on aneurysm screening programs, dietary improvements, and smoking reduction efforts. Additionally, in response to the challenges posed by an aging population, it is crucial to promote more comprehensive elderly health management services and enhance healthcare infrastructure. Furthermore, attention should be paid to the health protection of vulnerable populations, particularly in low-income countries and regions, by improving data collection and disease surveillance capacity to more accurately assess and address the disease burden. In developing countries, especially in Sub-Saharan Africa, strengthening basic healthcare infrastructure, improving healthcare accessibility, and fostering collaborations with international organizations and non-governmental organizations are essential to compensate for the deficiencies in public health services.

At the national level, we found that countries located closer to the poles exhibited higher mortality rates, while those near the equator showed the opposite trend. This phenomenon may be associated with the occurrence of abdominal AA in relation to colder temperatures. Research has demonstrated a correlation between lower temperatures and the incidence of aortic dissection, potentially due to the effects of cold weather on vascular walls and hypertension exacerbated by cold conditions^22, 23, 33-35^.

However, it is noteworthy that regional climate is merely one of many factors influencing mortality. Lifestyle choices, accessibility to healthcare services, and the quality of medical care also play critical roles. Therefore, in addition to climate factors, the combined impact of various socio-economic determinants on mortality should also be considered.

The relationship between DALY caused by AA and social demographic index (SDI) is not straightforward. The burden of AA is positively correlated with the level of national development, rising most rapidly in regions with medium SDI levels, while regions with high SDI levels exhibit the highest mortality rates, followed by a downward trend. Economic growth is typically accompanied by an increase in medical resources, making early diagnosis and treatment of aneurysms more feasible. Changes in lifestyle, such as the Westernization of dietary patterns and decreased physical activity, may contribute to the rising incidence of cardiovascular diseases^36^, including aneurysms, thereby increasing the number of disability-adjusted life years. Furthermore, advancements in medical technology have significantly improved the treatment outcomes for aneurysms, with increased success rates for surgical and interventional procedures^37^, as well as improvements in health management and chronic disease management. This indicates that there will be a substantial need for vascular surgery in medium to low SDI regions in the coming decades to reduce the mortality and related disability-adjusted life years associated with AA.

However, regional disparities and potential biases are crucial when interpreting these data. In high-SDI regions, despite the availability of ample healthcare resources and more opportunities for early diagnosis and treatment, lifestyle changes in high-income countries-such as unhealthy diets and physical inactivity-coupled with an aging population, may contribute to a sustained increase in the burden of aneurysms. This rising burden is not solely attributable to improvements in medical conditions but is also influenced by socio-economic, environmental factors, and health behaviors. In contrast, in low-income or middle-SDI regions, while healthcare resources may be insufficient and early diagnosis and treatment face considerable challenges, the burden of cardiovascular diseases may remain relatively lower, as lifestyle factors or dietary habits may differ.

At the same time, it is important to recognize that the Global Burden of Disease (GBD) data may also contain inherent biases. Firstly, due to variations in data collection and reporting, some low-income countries and regions may underestimate the burden of aneurysms, which is linked to the incomplete nature of health monitoring and medical records in these areas. Moreover, the Global Burden of Disease model relies on known diagnostic and treatment data, and in certain regions, healthcare standards and technologies may not have reached the level necessary to effectively diagnose and treat aneurysms, resulting in underreporting of cases. Therefore, comparisons of data across different countries and regions should be approached with caution, taking into account the disparities in healthcare systems and data reporting mechanisms.

The burden of AA peaks among the elderly. Physiologically, the aging population experiences a reduction in elastic fibers, an increase in collagen, lipid deposition, and inflammatory responses, all of which can lead to an increased risk of mortality^38–41^. Additionally, AA is associated with declining health status and various diseases, such as hypertension, and the natural increase in comorbidities among the elderly raises the mortality rate of AA. This study found that individuals aged 95 and older accounted for the highest number of AA-related deaths. Compared to never smokers, current smokers and former smokers have a 5-fold and 2-fold increased risk of developing AAA, respectively^42^. A positive dose-response relationship was observed between the number of cigarettes smoked daily and the pack-years of smoking^13^, indicating that older adults are more susceptible to the adverse effects of smoking.

Consistent with previous studies, we found that the mortality and DALY rates are slightly higher in men, primarily reflecting differences in smoking behaviors and occupational exposure to pollutants^43–45^. Research has shown that smoking is associated with nearly a 30-fold increased risk of AAA in women under 75 years old^46^. In our study, the mortality and DALY rates among women over 70 years old showed a narrowing gap compared to men, eventually surpassing men after the age of 90. This may be related to the decline in estrogen levels following menopause, which increases the risk of aneurysm formation in older women^47^. Investigating the role of estrogen therapy in preventing AA occurrence may become a focus of our future research.

In this study, smoking, high body-mass index and high systolic blood pressure were identified as the largest contributors to the burden of AA. These risk factors are largely preventable, and the high fatality rate of AA underscores the importance of prevention. Therefore, reducing the burden of AA necessitates targeted interventions and greater focus on mitigating these modifiable risks.

To address smoking as a major risk factor, in 2007, WHO endorsed a practical and cost-effective plan “MPOWER” for controlling the tobacco epidemic. This framework has been widely recognized as a practical and cost-effective approach to reducing smoking prevalence globally. In 2013, WHO further endorsed the Global Action Plan for the Prevention and Control of NCDs 2013-2020. This initiative emphasizes reducing tobacco use and promote sodium reduction in diets, support the development of hypertension screening and treatment programs in various countries, and enhance access to high-quality antihypertensive medications.^24^

Despite the global mean blood pressure appears to have stabilized due to the promotion of lifestyle modifications and pharmacological interventions, challenges still persist, such as hypertension awareness, treatment coverage, and control rates, particularly in low- and middle-income countries.

Building on these efforts, WHO released a new guideline for on the pharmacological treatment of hypertension in adults in 2021. The publication provides evidence-based recommendations for the initiation of treatment of hypertension, and recommended intervals for follow-up. The document also includes target blood pressure to be achieved for control, and information on who, in the health-care system, can initiate treatment.^25^

### Conclusions

AA remains an underrecognized and often fatal condition, frequently undiagnosed in the absence of adequate screening and diagnostic measures, thereby posing significant challenges for its treatment and management. Our study demonstrates that by enhancing screening efforts and controlling risk factors associated with AA, mortality and DALYs can be effectively reduced. However, given the unequal distribution of resources across different global regions and the exacerbating trend of population aging, the burden of AA is expected to rise further. These findings offer valuable guidance to policymakers, particularly public health leaders in low- and middle-income countries, assisting them in advancing more comprehensive vascular health initiatives to mitigate the public health burden associated with AA.

## Contributors

FHH and WC contributed equally as corresponding authors and designed the study. TJ and QL analysed the data and performed the statistical analyses. TJ, QL, DLC, XLZ, YYZ, JL, SCL, WC and FHH drafted the initial manuscript. FHH and WC are the guarantors. All authors reviewed the drafted manuscript for critical content. All authors approved the final version of the manuscript. The corresponding authors (FHH and WC) attest that all listed authors meet authorship criteria and that no others meeting the criteria have been omitted.

## Ethical approval

This study utilized exclusively aggregated, de-identified data from the Global Burden of Disease (GBD) database, which is publicly accessible and complies with ethical guidelines for secondary data analysis. Formal ethical approval and informed consent were not required for this research. All analyses were conducted in accordance with the GBD Data Use Agreement.

## Data sharing

The data used for the analyses in the study are publicly available at https://ghdx.healthdata.org/gbd-results-tool.

## Dissemination to participants and related patient and public communities

The results will be disseminated through media outlets and presentations at scientific conferences and academic events. Given that no patients were recruited for the study, there are no plans to disseminate the results to study participants.

The manuscript’s guarantors (FHH and WC) affirm that the manuscript is an honest, accurate, and transparent account of the study being reported; that no important aspects of the study have been omitted; and that any discrepancies from the study as planned (and, if relevant, registered) have been explained.

Provenance and peer review: Not commissioned; externally peer reviewed.

## Figure Legends

**Fig 1.** Age standardised death rate of aortic aneurysm per 100 000 population in 2021, by country (generated from data available at https://ghdx.healthdata.org/gbd-results-tool)

**Fig2** Age standardised disability adjusted life year (DALY) rate of aortic aneurysm per 100 000 population in 2021, by country (generated from data available at https://ghdx.healthdata.org/gbd-results-tool)

**Fig 3** Age standardised disability adjusted life year (DALY) rates of aortic aneurysm for the 21 Global Burden of Disease regions by sociodemographic index, 1990–2021. Thirty points are plotted for each region and show the observed age standardised DALY rates from 1990 to 2021 for that region. Expected values, based on sociodemographic index and disease rates in all locations, are shown as a solid line. Regions above the solid line represent a higher than expected burden (eg, Oceania) and regions below the line show a lower than expected burden (eg, East Asia) (generated from data available at https://ghdx.healthdata.org/gbd-results-tool)

**Fig 4** Percentage of disability adjusted life years (DALYs) due to aortic aneurysm attributable to each risk factor for the 21 Global Burden of Disease regions in 2021 (generated from data available at https://ghdx.healthdata.org/gbd-results-tool)

**Fig 5** Percentage of disability adjusted life years (DalYs) due to aortic aneurysm attributable to each risk factor for the 21 global burden of Disease regions in 2021 (generated from data available at https://ghdx.healthdata.org/gbd-results-tool)

**Figure S1:** Age standardised death rate of aortic aneurysm per 100 000 population in 2021 for the 55 Global Burden of Disease regions, by sex. (Generated from data available from http://ghdx.healthdata.org/gbd-results-tool).

**Figure S2:** Age standardised DALY rates of aortic aneurysm in 2021 for the 55 Global Burden of Disease regions, by sex. DALY=disability adjusted life years. (Generated from data available from http://ghdx.healthdata.org/gbd-results-tool).

**Figure S3:** The percentage change in the age-standardised death rates of aortic aneurysm from 1990 to 2021 for the 55 Global Burden of Disease regions, by sex. (Generated from data available from http://ghdx.healthdata.org/gbd-results-tool).

**Figure S4:** The percentage change in the age-standardised DALY rates of aortic aneurysm from 1990 to 2021 for the 55 Global Burden of Disease regions, by sex. DALY=disability adjusted life years. (Generated from data available from http://ghdx.healthdata.org/gbd-results-tool).

**Figure S5:** Global number of DALYs and DALY rate of aortic aneurysm per 100,000 population, by age and sex, in 2021; Dotted and dashed lines indicate 95% upper and lower uncertainty intervals, respectively. DALY=disability adjusted life years. (Generated from data available from http://ghdx.healthdata.org/gbd-results-tool).

**Figure S6:** Age-standardised DALY rates of aortic aneurysm for 204 countries and territories, by SDI, in 2021; Expected values based on the Socio-demographic Index and disease rates in all locations are shown as the black line. Each point shows the observed age-standardised DALY rate for each country in 2021. DALY=disability adjusted life years. SDI= Socio-demographic Index (Generated from data available from http://ghdx.healthdata.org/gbd-results-tool).

**Figure S7:** Percentage of DALYs due to aortic aneurysm attributable to risk factors among males for 21 GBD regions in 2021. DALY=disability adjusted life years (Generated from data available from, http://ghdx.healthdata.org/gbd-results-tool).

**Figure S8:** Percentage of DALYs due to aortic aneurysm attributable to risk factors among females for 21 GBD regions in 2021. DALY=disability adjusted life years (Generated from data available from http://ghdx.healthdata.org/gbd-results-tool).

**Figure S9:** Percentage of DALYs due to aortic aneurysm attributable to each risk factor, by age, in 2021. DALY=disability adjusted life years (Generated from data available from http://ghdx.healthdata.org/gbd-results-tool).

**Figure S10:** Percentage of DALYs due to aortic aneurysm attributable to each risk factor among males, by age, in 2021. DALY=disability adjusted life years (Generated from data available from http://ghdx.healthdata.org/gbd-results-tool).

**Figure S11:** Percentage of DALYs due to aortic aneurysm attributable to each risk factor among females, by age, in 2021. DALY=disability adjusted life years (Generated from data available from http://ghdx.healthdata.org/gbd-results-tool).

## Uncategorized References

1. Quintana RA and Taylor WR. Cellular Mechanisms of Aortic Aneurysm Formation. Circ Res. 2019;124:607–618.

2. Golledge J. Abdominal aortic aneurysm: update on pathogenesis and medical treatments. Nat Rev Cardiol. 2019;16:225–242.

3. Nordon IM, Hinchliffe RJ, Loftus IM and Thompson MM. Pathophysiology and epidemiology of abdominal aortic aneurysms. Nat Rev Cardiol. 2011;8:92–102.

4. Bossone E and Eagle KA. Epidemiology and management of aortic disease: aortic aneurysms and acute aortic syndromes. Nat Rev Cardiol. 2021;18:331–348.

5. Wang Z, You Y, Yin Z, Bao Q, Lei S, Yu J, Xie C, Ye F and Xie X. Burden of Aortic Aneurysm and Its Attributable Risk Factors from 1990 to 2019: An Analysis of the Global Burden of Disease Study 2019. Front Cardiovasc Med. 2022;9:901225.

6. Huynh TT, Miller CC, 3rd, Estrera AL, Porat EE and Safi HJ. Thoracoabdominal and descending thoracic aortic aneurysm surgery in patients aged 79 years or older. J Vasc Surg. 2002;36:469–75.

7. Jahangir E, Lipworth L, Edwards TL, Kabagambe EK, Mumma MT, Mensah GA, Fazio S, Blot WJ and Sampson UK. Smoking, sex, risk factors and abdominal aortic aneurysms: a prospective study of 18 782 persons aged above 65 years in the Southern Community Cohort Study. J Epidemiol Community Health. 2015;69:481–8.

8. Mitsouras D and Leach JR. Expanding the Radiologist’s Arsenal against Abdominal Aortic Aneurysms, a Versatile Adversary. Radiology. 2020;295:730–732.

9. Dua A, Kuy S, Lee CJ, Upchurch GR, Jr. and Desai SS. Epidemiology of aortic aneurysm repair in the United States from 2000 to 2010. J Vasc Surg. 2014;59:1512–7.

10. Kim HW and Stansfield B. Genetic and Epigenetic Regulation of Aortic Aneurysms. BioMed Research International. 2015;2017:1–12.

11. Zhou J, Lin J and Zheng Y. Association of cardiovascular risk factors and lifestyle behaviors with aortic aneurysm: A Mendelian randomization study. Front Genet. 2022;13.

12. Svensjö S, Mani K, Björck M, Lundkvist J and Wanhainen A. Screening for Abdominal Aortic Aneurysm in 65-Year-old Men Remains Cost-effective with Contemporary Epidemiology and Management. European Journal of Vascular and Endovascular Surgery. 2014;47:357–365.

13. Aune D, Schlesinger S, Norat T and Riboli E. Tobacco smoking and the risk of abdominal aortic aneurysm: a systematic review and meta-analysis of prospective studies. Sci Rep. 2018;8:14786.

14. Huang H, Tang L, Liu C and Jin G. Trends and risk factors analysis of aortic aneurysm mortality in China over thirty years: based on the global burden of disease 2019 data. Eur Heart J Qual Care Clin Outcomes. 2024:qcae084.

15. Wang H, Yu X, Guo J, Ma S, Liu Y, Hu Y, Li J, Song Y and Zou Z. Burden of cardiovascular disease among the Western Pacific region and its association with human resources for health, 1990-2021: a systematic analysis of the Global Burden of Disease Study 2021. Lancet Reg Health West Pac. 2024;51:101195.

16. Wei L, Bu X, Wang X, Liu J, Ma A and Wang T. Global Burden of Aortic Aneurysm and Attributable Risk Factors from 1990 to 2017. Glob Heart. 2021;16:35.

17. Naghavi M, Ong KL, Aali A, Ababneh HS, et al. Global burden of 288 causes of death and life expectancy decomposition in 204 countries and territories and 811 subnational locations, 1990–2021: a systematic analysis for the Global Burden of Disease Study 2021. The Lancet. 2024;403:2100–2132.

18. Jazieh AR, Akbulut H, Curigliano G, Rogado A, Alsharm AA, Razis ED, Mula-Hussain L, Errihani H, Khattak A, De Guzman RB, Mathias C, Alkaiyat MOF, Jradi H, Rolfo C and International Research Network on C-IoCC. Impact of the COVID-19 Pandemic on Cancer Care: A Global Collaborative Study. JCO Global Oncology. 2020;6:1428–1438.

19. Jiang D, Wu Y, Liu L, Shen Y, Li T, Lu Y, Wang P, Sun C, Wang K, Wang K and Ye H. Burden of Gastrointestinal Tumors in Asian Countries, 1990–2021: An Analysis for the Global Burden of Disease Study 2021. CLEP. 2024;Volume 16:587–601.

20. Png CYM, Wu J, Tang TY, Png IPL, Sheng TJ and Choke E. Editor’s Choice - Decrease in Mortality from Abdominal Aortic Aneurysms (2001 to 2015): Is it Decreasing Even Faster? Eur J Vasc Endovasc Surg. 2021;61:900–907.

21. Diseases GBD and Injuries C. Global incidence, prevalence, years lived with disability (YLDs), disability-adjusted life-years (DALYs), and healthy life expectancy (HALE) for 371 diseases and injuries in 204 countries and territories and 811 subnational locations, 1990-2021: a systematic analysis for the Global Burden of Disease Study 2021. Lancet. 2024;403:2133–2161.

22. Choong A, Marjot J, Wee IJY, Syn N, Marjot T, Brightwell RE and Walker PJ. Forecasting aortic aneurysm rupture: A systematic review of seasonal and atmospheric associations. J Vasc Surg. 2019;69:1615–1632 e17.

23. Yu X, Xia L, Xiao J, Zheng J, Xu N, Feng X and Wei X. Association of Daily Mean Temperature and Temperature Variability With Onset Risks of Acute Aortic Dissection. J Am Heart Assoc. 2021;10:e020190.

24. (NCD) WTNDRaD. Global action plan for the prevention and control of noncommunicable diseases 2013-2020. 2013.

25. Guideline for the pharmacological treatment of hypertension in adults. Geneva: World Health Organization; 2021. Licence: CC BY-NC-SA 3.0 IGO.

26. Mills KT, Bundy JD, Kelly TN, Reed JE, Kearney PM, Reynolds K, Chen J and He J. Global Disparities of Hypertension Prevalence and Control: A Systematic Analysis of Population-Based Studies From 90 Countries. Circulation. 2016;134:441–50.

27. Collaboration NCDRF. Worldwide trends in blood pressure from 1975 to 2015: a pooled analysis of 1479 population-based measurement studies with 19.1 million participants. Lancet. 2017;389:37–55.

28. Collaborators GBDT. Spatial, temporal, and demographic patterns in prevalence of smoking tobacco use and attributable disease burden in 204 countries and territories, 1990-2019: a systematic analysis from the Global Burden of Disease Study 2019. Lancet. 2021;397:2337–2360.

29. Collaborators GBDCT. Spatial, temporal, and demographic patterns in prevalence of chewing tobacco use in 204 countries and territories, 1990-2019: a systematic analysis from the Global Burden of Disease Study 2019. Lancet Public Health. 2021;6:e482–e499.

30. de Boer AR, Vaartjes I, van Dis I, van Herwaarden JA, Nathoe HM, Ruigrok YM, Bots ML, Visseren FLJ and group U-Ss. Screening for abdominal aortic aneurysm in patients with clinically manifest vascular disease. Eur J Prev Cardiol. 2022;29:1170–1176.

31. Guirguis-Blake JM, Beil TL, Senger CA and Coppola EL. Primary Care Screening for Abdominal Aortic Aneurysm: Updated Evidence Report and Systematic Review for the US Preventive Services Task Force. JAMA. 2019;322:2219–2238.

32. Sprynger M, Willems M, Van Damme H, Drieghe B, Wautrecht JC and Moonen M. Screening Program of Abdominal Aortic Aneurysm. Angiology. 2019;70:407–413.

33. Huang HN, Li X, Peng Z, Liao YF, Li L, Nardocci AC, Ou CQ and Yang Z. Mortality risk and burden of aortic aneurysm and dissection attributable to low temperatures: A nationwide case-crossover analysis in Brazil, a predominantly tropical country. Environ Int. 2024;190:108895.

34. Ma WG, Li B, Zhang W, Sarkar AEH, Tufail R, Peterss S, Zheng J, Zafar MA, Ziganshin BA, Zhu JM, Sun LZ and Elefteriades JA. Chronologic and Climatic Factors of Acute Aortic Dissection: Study of 1642 Patients in Two Continents. Ann Thorac Surg. 2020;110:575–581.

35. Taheri Shahraiyni H, Sodoudi S and Cubasch U. Weather conditions and their effect on the increase of the risk of type A acute aortic dissection onset in Berlin. Int J Biometeorol. 2016;60:1303–5.

36. Wu G, Zhang X and Gao F. The epigenetic landscape of exercise in cardiac health and disease. J Sport Health Sci. 2021;10:648–659.

37. Schmitz-Rixen T, Bockler D, Vogl TJ and Grundmann RT. Endovascular and Open Repair of Abdominal Aortic Aneurysm. Dtsch Arztebl Int. 2020;117:813–819.

38. Feng Y, Wang X, Zhao Y, Li L, Niu P, Huang Y, Han Y, Tan W and Huo Y. A comparison of passive and active wall mechanics between elastic and muscular arteries of juvenile and adult rats. J Biomech. 2021;126:110642.

39. Samouillan V, Dandurand J, Lacabanne C, Stella A, Gargiulo M, Degani A, Gandaglia A and Spina M. Analysis of the molecular mobility of collagen and elastin in safe, atheromatous and aneurysmal aortas. Pathol Biol (Paris). 2012;60:58–65.

40. Chao de la Barca JM, Richard A, Robert P, Eid M, Fouquet O, Tessier L, Wetterwald C, Faure J, Fassot C, Henrion D, Reynier P and Loufrani L. Metabolomic Profiling of Angiotensin-II-Induced Abdominal Aortic Aneurysm in Ldlr(-/-) Mice Points to Alteration of Nitric Oxide, Lipid, and Energy Metabolisms. Int J Mol Sci. 2022;23.

41. Yu H, Jiao X, Lv Q, Li L, Du Y, Hu C, Du Z, Li F, Wang Y, Gao X, Han L, Sun X, Chen D and Qin Y. ATF4 Contributes to Abdominal Aortic Aneurysm Formation via Modulating M1 Macrophage Polarization and Inflammation. Aging Dis. 2024.

42. Sode BF, Nordestgaard BG, Gronbaek M and Dahl M. Tobacco smoking and aortic aneurysm: two population-based studies. Int J Cardiol. 2013;167:2271–7.

43. Sweeting MJ, Thompson SG, Brown LC, Powell JT and collaborators R. Meta-analysis of individual patient data to examine factors affecting growth and rupture of small abdominal aortic aneurysms. Br J Surg. 2012;99:655–65.

44. Fagerberg B, Borne Y, Sallsten G, Smith JG, Acosta S, Persson M, Melander O, Forsgard N, Gottsater A, Hedblad B, Barregard L and Engstrom G. Circulating cadmium concentration and risk of aortic aneurysms: A nested case-control study within the Malmo Diet and Cancer cohort. Atherosclerosis. 2017;261:37–43.

45. Kerns E, Masterson EA, Themann CL and Calvert GM. Cardiovascular conditions, hearing difficulty, and occupational noise exposure within US industries and occupations. Am J Ind Med. 2018;61:477–491.

46. Carter JL, Morris DR, Sherliker P, Clack R, Lam KBH, Halliday A, Clarke R, Lewington S and Bulbulia R. Sex-Specific Associations of Vascular Risk Factors With Abdominal Aortic Aneurysm: Findings From 1.5 Million Women and 0.8 Million Men in the United States and United Kingdom. J Am Heart Assoc. 2020;9:e014748.

47. Egorova NN, Vouyouka AG, McKinsey JF, Faries PL, Kent KC, Moskowitz AJ and Gelijns A. Effect of gender on long-term survival after abdominal aortic aneurysm repair based on results from the Medicare national database. J Vasc Surg. 2011;54:1–12 e6; discussion 11-2.

